# Usages and perceptions of artificial intelligence among French radiologists

**DOI:** 10.64898/2026.03.23.26348621

**Authors:** Aurélie Jean, Thibaut Jacques, Philippe Benillouche

## Abstract

This study analyzes the adoption, barriers, and expectations of French radiologists regarding the use of Artificial Intelligence (AI) solutions in their daily practice. Despite a recognition of AI’s potential to make radiology more precise, predictive, and personalized, its adoption remains limited. The main obstacles identified are the high cost of those solutions and the insufficient equipment of French imaging centers with AI technologies. Nevertheless, the survey reveals a strong willingness to adopt, with over 70% of radiologists expressing their desire to use AI and 0% declaring a refusal to use it. Furthermore, the radiologists’ fears of being replaced by AI are very low (0 to 8.8%).

## INTRODUCTION: CONTEXT AND CHALLENGES

Initially used in radiology for segmentation and classification of medical images, AI now plays a role in aiding diagnosis (1). The integration of AI in diagnostic radiology promises to improve the interpretation of medical images (2)(3) through the detection of subtle or weak signals (precision), the anticipation of prognoses (prediction), and the adaptation of patient follow-ups (personalization). This allows medicine to enter a new preventive paradigm that combines greater precision, prediction, and personalization (4)(5)(6). This is the case for diagnostic breast imaging (7)(8)(9), which could help improve breast cancer screening campaigns (10).

In France, although the integration of AI in radiology is progressing, its deployment remains heterogeneous and is more widespread in University Hospitals (11) where 80% of surveyed radiologists report using it. According to the *Institut Curie* in 2024 (12), mammography appears to be the most advanced domain in radiology in terms of adoption and integration of AI for image interpretation. In 2023 (13), 10% of radiology centers were estimated to be equipped with AI for mammographic image interpretation. It is thus estimated that 20 to 30% of radiologists currently use a diagnostic AI solution in mammography.

To date, there are no precise figures on the actual adoption of AI in aid of medical image interpretation in radiology in France. The *Société Française de Radiologie*, particularly through *DRIM France IA*, maintains a scientific and technological watch. *DRIM France IA* (14) recently crossed a major milestone by publishing independent evaluation grids for tools available on the market, especially for AI solutions in mammography. These grids give the structures the possibility to compare AI solutions not only based on their standardized performance but also regarding the human guarantees they offer.

The survey presented here aims to understand the reasons for still limited adoption and to identify levers to accelerate the integration of AI into the daily practice of medical image interpretation by radiologists. In 2019, a similar study, led by Dr. Jacques (15), was conducted on the perception of AI by French radiologists, highlighting their appetite for adopting AI in image interpretation, as well as their desire for more training. A study conducted in 2024 (16) jointly by the *Docteur Imago media and Tech Imago* on 144 medical imaging professionals, including 75 radiologists, shows that 81% of them, across all disciplines, use AI, with 77% using it regularly for all use cases such as diagnosis, image quality improvement, contouring, or report writing assistance.

## METHODOLOGY

### Study description and radiologist recruitment

The study was conducted with a limited sample of 100 radiologists practicing throughout France, including the overseas departments and territories (DOM-TOM). Recruitment was carried out *via* professional platforms (*LinkedIn*), professional networks, and the solicitation of groups of radiology centers.

*Table 1* presents the geographic origin of the radiologists’ activity. The overrepresentation of radiologists in Île-de-France (48%) is notable, aligning with national demographic data (DREES 2025) (17). In practice, data from the *Direction de la Recherche, des Études, de l’Évaluation et des Statistiques* show a density of 18 to 20 radiologists per 100,000 inhabitants in the Paris region (reaching up to 40 per 100,000 in Paris), a strong contrast with the 11 to 13 radiologists in other territories and the rate below 6 per 100,000 in certain areas considered critical.

**Table 1:** Geographic origins of the professional activity of responding radiologists.

| Region |  |  |  |
| --- | --- | --- | --- |
| Auvergne-Rhône-Alpes | 8% | Nouvelle-Aquitaine | 3% |
| Bourgogne-Franche-Comté | 2% | Occitanie | 8% |
| Bretagne | 4% | Pays de la Loire | <1% |
| Centre-Val de Loire | 8% | Provence-Alpes-Côte d'Azur | 3% |
| Corse | <1% | Guadeloupe | <1% |
| Grand Est | 4% | Guyane | <1% |
| Hauts-de-France | 9% | Martinique | <1% |
| Île-de-France | 48% | Mayotte | <1% |
| Normandie | 1% |  |  |

The study sample includes experts, with 72% of them interpreting mammograms. *Table 2* presents their distribution according to the number of years of practice. The largest portion of radiologists here have between 20 and 30 years of experience. The percentage of experts reaches 89% if all those with more than 10 years of experience are considered. These figures align with DREES data, which places the average age of radiologists above 50, and highlights an aging population of practicing radiologists in France, rising from 7.2% of radiologists over 65 in 2012 to 18.7% in 2023. *Table 3* presents the average number of mammograms interpreted per month by the responding radiologists. These figures are aligned with the specifications from the organized screening protocol, which requires a minimum of 500 mammograms read per year during the first reading by radiologists integrated into the program (source: Ameli (18)).

**Table 2:** Number of years in practice for the 72% of responding radiologists interpreting mammographic images.

| Nombre d'année d'exercice en radiologie mammaire |  |  |  |
| --- | --- | --- | --- |
| Less than 5 years | 6.9% | Between 20 and 30 years | 38.9% |
| Between 5 and 10 years | 4.2% | More than 30 years | 22.2% |
| Between 10 and 20 years | 27.8% |  |  |

**Table 3:** Monthly volume of mammograms interpreted by the 72% of responding radiologists interpreting mammograms.

| Monthly volume of mammography readings |  |  |  |
| --- | --- | --- | --- |
| Less than 100 | 18.1% | Between 500 and 750 | 8.3% |
| Between 100 and 250 | 37.5% | More than 750 | 9.7% |
| Between 250 and 500 | 26.4% |  |  |

### Data Collection

Participants responded to an online *Google* questionnaire (*Google Form*) compliant with the General Data Protection Regulation (GDPR). The questionnaire consisted of 10 questions distinguishing radiologists who use AI from those who do not, as well as radiologists who practice mammography daily from those who do not.

Among these questions, those concerning the respondents’ profile were mandatory (number of years of practice, region of practice, volume of readings, interpreting - or not - mammographic images, and use of AI in radiological image interpretation). The other questions were made optional to avoid skewing the responses by blocking or forcing participants from an uncontrolled (i.e., not predefined) sample to respond.

### Data Analysis

Results were analyzed and presented as percentages, based on the pre-analysis performed by *Google* tools and the database storing all the results.

## RESULTS

**Survey Results**

The main results of the survey are presented here, distinguishing the responses of radiologists who interpret mammograms from those of other radiologists. Systematically, the number of responses provided is associated with the percentage calculated based on the number of respondents to each question.

### Barriers to Use (*Table 5*)

- Among mammography experts not using AI:
  ○ The main obstacle is a lack of equipment for 94.1% of them,
  ○ The cost is a limiting factor for 35.3% of them,
  ○ 8.8% express a lack of confidence in AI solutions.
- Among other radiologists not using AI:
  ○ 71.4% are not equipped,
  ○ 28.6% mention cost and lack of confidence,
  ○ 42.9% do not perceive any added value of AI in their practice.

### Workflow Improvement (*Table 7*)

Among radiologists using AI in their daily practice of radiological image interpretation:

- 65.8% of radiologists reading mammograms report seeing their workflow improved, and 5.3% report not knowing the answer,
- 85.7% of other radiologists report seeing their workflow improved, and 4.8% report not knowing.

### Adoption Intentions and Fears (*Tables 8 and 9*)

- A very strong majority of surveyed radiologists, who are not yet using AI, wish to adopt it: 79.4% of mammography experts and 71.4% of other radiologists (*Table 8*),
- Remarkably: 0% of respondents decline the idea of using AI (*Table 8*),
- Furthermore, the fear of being replaced by AI in the long term is extremely low, cited by only 8.8% of mammography experts and 0% of other radiologists (*Table 7*).

### Radiologists’ Expectations (*Table 10*)

Among the radiologists’ responses concerning their expectations for AI solutions, three return most frequently:

- *Aid to diagnosis*,
- *Reduction of errors* (false negatives and false positives),
- *Time saving and productivity improvement*.

*Aid to diagnosis* is a more marked expectation among radiologists interpreting mammograms who use (46.2%) or do not use (44%) AI. *Aid to diagnosis* and *Time saving* are more marked expectations among other radiologists who use (29.4% and 23.5%) or do not use (28.7% and 48.9%) AI.

## DISCUSSION

### Discussion of the sample

The sample of the study, considered here, remains small compared to the samples of other studies, particularly the one from 2019 (15), which included 270 responding radiologists. The study conducted in 2024 jointly by the *Docteur Imago* media and *Tech Imago* (16) was composed of 144 medical imaging professionals, including 75 radiologists, 46 manipulators, and 14 health executives. Given the size of the sample considered here (100 radiologists), particular attention must be paid to the analysis of results and the risks of bias.

*Table 4* presents the proportions of radiologists using AI in radiological image interpretation. These figures are higher than the national statistics (20 to 30% in mammography compared to the 52.8% obtained here). This can be explained by a selection bias among respondents, as those who already use AI for image interpretation are more inclined to respond to this questionnaire. Also, among the respondents, the proportion of non-breast-imaging radiologists using AI (75%) is higher than that of radiologists interpreting mammograms (52.8%). This variability between sub-specialties of radiology is found in the literature (19). One of the main factors could be the lower proportion of CE-marked algorithms specialized in breast imaging: about 10% of all algorithms, compared to 90% for the rest of radiology (20), thus facilitating wider deployment to other sub-specialties. Other issues such as integration into the workflow of radiologists, and managing discrepancies can be mentioned.

**Table 4:** Use of AI in daily practice in radiology by responding radiologists.

| Use of AI in the daily practice of image interpretation<br>(number and percentage) |  |  |  |  |
| --- | --- | --- | --- | --- |
|  | Among radiologists interpreting mammograms |  | Among other radiologists |  |
| YES | 38 | 52.8% | 21 | 75% |
| NO | 34 | 47.2% | 7 | 25% |

**Table 5:**
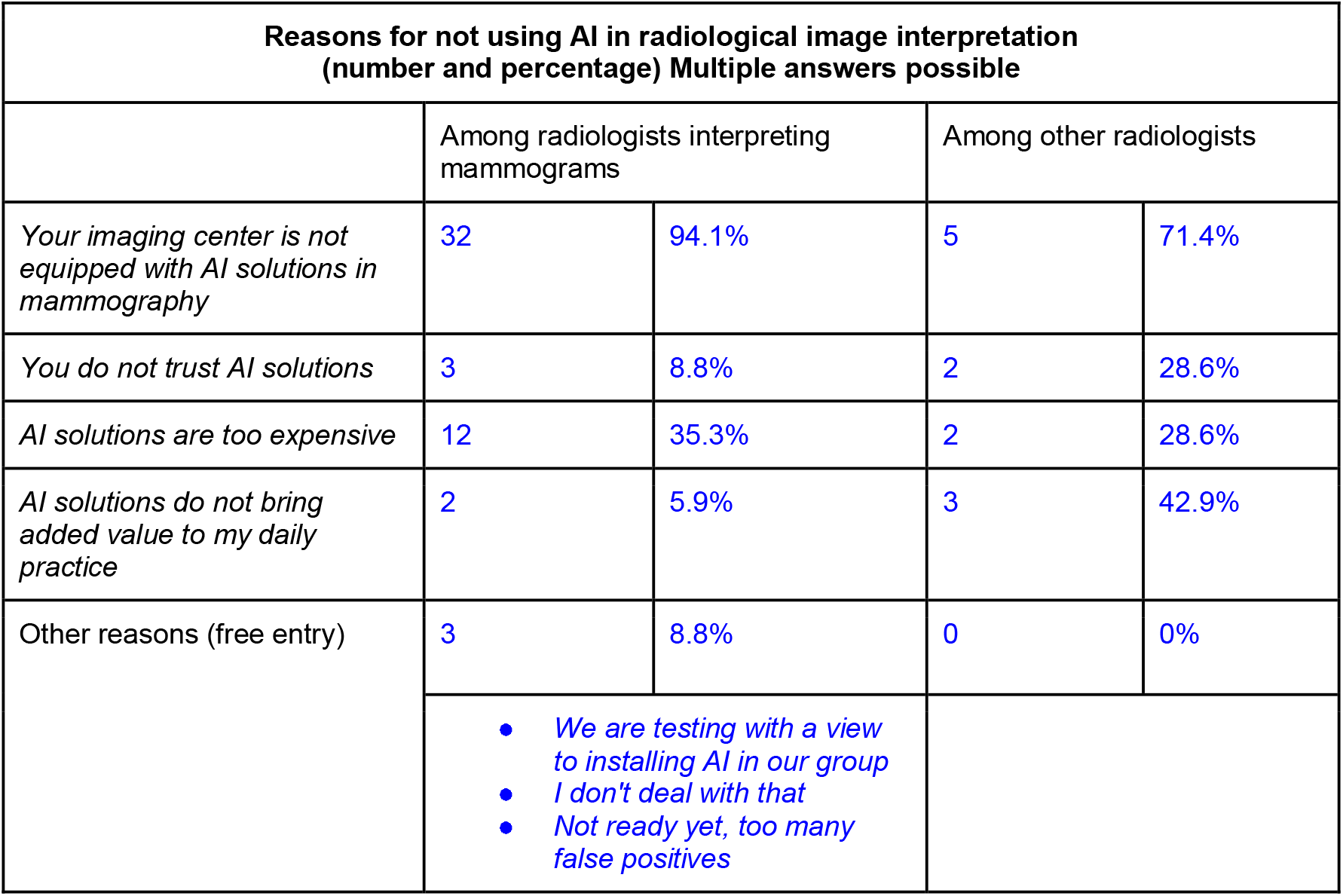
Reasons for not using AI in daily practice in radiology by responding radiologists who currently do not use any AI in image interpretation.

The sample (see *Table 1*) has an overrepresentation of respondents practicing in Île-de-France (48%) compared to 20 to 25% of doctors (all disciplines combined) practicing in France who are concentrated in the Paris region (sources: *Atlas of Medical Demography 2025* by the National Council of the Order of Physicians (21) and *Demographic Analysis of Radiologists in France since 2012* (22)). This overrepresentation is notably explained by the extent of the radiologists’ networks that the survey authors were able to solicit, which are more present in this region.

The sample has a lower proportion of respondents with less than 10 years (10.11%), including less than 5 years (6.9%) of experience (see *Table 6*). This difference can be explained by the lack of time of young radiologists (and especially residents) to respond to this type of survey.

**Table 6:** Number of years of AI use in image interpretation among responding radiologists.

| Number of years of AI use in radiology<br>(number and percentage) |
| --- |

|  | Among radiologists interpreting mammograms |  | Among other radiologists |  |
| --- | --- | --- | --- | --- |
| Less than 2 years | 14 | 36.8% | 4 | 19% |
| Between 2 and 5 years | 12 | 31.6% | 13 | 61.9% |
| Between 5 and 10 years | 8 | 21.1% | 4 | 19% |
| More than 10 years | 4 | 10.5% | 0 | 0% |

### Discussion of the results

The results of this survey confirm that radiologists are ready to adopt AI. The analysis indicates that nearly 8 out of 10 mammography experts who do not yet use AI are ready to integrate it immediately, even if 42.9% of them do not yet clearly perceive the added value of AI in their practice. None report refusing to use an AI solution if it were offered to them for free. This highlights their appetite for these tools, which are still considered inaccessible by 35.3% of radiologists in mammography and 23.6% of other radiologists, due to their estimated high costs. Their expectations are high and legitimate. Their concerns focus less on the fear of replacement than on pragmatic factors such as cost and equipment.

*Table 7* presents the proportions of radiologists stating that AI improves their workflow. Breast imaging specialists are less numerous (65.8%) to state that AI improves their workflow, compared to other radiologists (85.7%). These data align with a recent study (23) where only 41.2% of breast imaging specialists reported an improvement in their workflow, with the rest of the panel being negatively impacted by increased time spent deliberating on doubtful cases and discrepancies. A literature review on the limitations of AI adoption in breast imaging actually reports a multifactorial origin (24): besides technological requirements and the need for reproducible clinical evidence, integration faces the challenge of trust and transparency, legal and ethical voids, as well as persistent uncertainties about its long-term societal impact. The fact that breast imaging specialists adopt AI less and report a smaller improvement in their workflow highlights the need to develop effective and relevant AI tools for mammogram interpretation.

**Table 7:** Proportions of responding radiologists, who use AI in radiological image interpretation, who state that it improves their workflow.

| Workflow improvement among radiologists using AI in image interpretation (number and percentage) |  |  |  |  |
| --- | --- | --- | --- | --- |
|  | Among radiologists interpreting mammograms |  | Among other radiologists |  |
| YES | 25 | 65.8% | 18 | 85.7% |
| NO | 11 | 28.9% | 2 | 9.5% |
| I don't know | 2 | 5.3% | 1 | 4.8% |

**Table 8:** Proportions of responding radiologists, who use and who do not use AI in radiological image interpretation, who state that it improves their workflow.

| Fear of being replaced by AI among radiologists using AI in image interpretation (number and percentage) |  |  |  |  |
| --- | --- | --- | --- | --- |
|  | Among radiologists interpreting mammograms |  | Among other radiologists |  |
| YES | 4 | 10.5% | 2 | 9.5% |
| NO | 31 | 81.6% | 17 | 80.1% |
| I don't know | 3 | 7.9% | 1 | 9.5% |
| Fear of being replaced by AI among radiologists not using AI in image interpretation (number and percentage) |  |  |  |  |
|  | Among radiologists interpreting mammograms |  | Chez les autres radiologues |  |
| YES | 3 | 8.8% | 0 | 0% |
| NO | 27 | 79.4% | 6 | 85.7% |
| I don't know | 4 | 11.8% | 1 | 14.3% |

**Table 9:** Proportions of responding radiologists, who do not use AI in radiological image interpretation, who state that they wish to have free access to an AI solution.

| **Desire among radiologists not using AI in image interpretation to have an AI solution available for free (number and percentage)** |  |  |  |  |
| --- | --- | --- | --- | --- |
|  | Among radiologists interpreting mammograms |  | Among other radiologists |  |
| YES | 27 | 79.4% | 5 | 71.4% |
| NO | 0 | 0% | 0 | 0% |
| I don't know | 7 | 20.6% | 2 | 28.6% |

**Table 10:** Main responses entered in the free text field by responding radiologists regarding their expectations for AI solutions in radiological image interpretation.

| **Main expectations of radiologists (free entry) using AI in image interpretation (number and percentage)** |  |  |  |  |
| --- | --- | --- | --- | --- |
|  | Among radiologists interpreting mammographic images |  | Among other radiologists |  |
| *Aid to diagnosis* | 7 | 46.2% | 5 | 29.4% |
| *Reduction of errors* | 4 | 30.8% | 3 | 17.6% |
| *Time saving / Productivity improvement* | 0 | 0% | 4 | 23.5% |
| **Main expectations of radiologists (free entry) not using AI in image interpretation (number and percentage)** |  |  |  |  |
|  | Among radiologists interpreting mammographic images |  | Among other radiologists |  |
| *Aid to diagnosis* | 11 | 44% | 2 | 28.7% |
| *Reduction of errors* | 6 | 24% | 0 | 0% |
| *Time saving / Productivity improvement* | 6 | 24% | 3 | 42.9% |

In general, radiologists demand to be reassured and to understand the AI tools they use. It is important, even critical, to implement methods to build a foundation of trust between the radiologist and the AI solution designer. To reassure physicians about the relevance of an AI tool and allow them to contest its results with full knowledge, its owner must guarantee, among other things, transparent communication about its governance. This governance includes good development practices, tests performed, and statistical performance results in different usage scenarios. To this governance, a regulatory framework such as the European text, the AI Act, is added, which implies a minimum of explainability and interpretability of AI tools.

To encourage adoption by radiologists, it is essential to propose solutions that directly meet their expectations, integrate into their workflow to increase their productivity, and are understandable. Furthermore, it is fundamental to support radiologists in their adoption of AI solutions by providing them with adequate and effective training content.

## CONCLUSION

The integration of AI in radiology represents a powerful lever to improve diagnoses and move towards greater precision, prediction, and personalization for the benefit of patients. The deployment of AI solutions, still limited due to excessive financial cost, needs to be facilitated while supporting radiologists in adopting AI solutions that they are already ready to embrace.

## Data Availability

All data produced in the present work are contained in the manuscript

## ACKNOWLEDGMENTS

The authors thank all the participants in this study for agreeing to respond to the survey. The authors also thank all the radiologists, radiology centers, hospital centers, as well as professional health networks that agreed to share the survey with their colleagues and community.

## AUTHOR CONTRIBUTIONS

Dr. Aurélie Jean (Ph.D.) and Dr. Philippe Benillouche (M.D.) contributed to the development and analysis of the survey. Dr. Thibaut Jacques (M.D., M.Sc.) contributed to the writing of this article and the discussion of the results.

